# Aligning Definitions with Realities: An Interpretive Descriptive Study on the Complexities of Measuring Retention in HIV Care in the Global Context

**DOI:** 10.64898/2026.02.13.26345822

**Authors:** Nadia Rehman, Gordon Guyatt, Lora Sabin, Juana Xiong, Mahala G. English, Gabriella Moraes Rae, Jessica E. Haberer, Michael Mugavero, Thomas P. Giordano, Dominik Mertz, Aaron Jones

## Abstract

**Background:** Retention in HIV care is associated with higher rates of antiretroviral treatment adherence and viral suppression, as well as lower risk of AIDS-related morbidity and mortality. However, the multidimensional nature of retention complicates measurement standardization, limiting comparability and global evaluation. This study explored how HIV stakeholders define and assess retention, aiming to develop a patient-centred and conceptually robust understanding to inform research and practice.

**Methods:** We conducted a qualitative study using Interpretive Description (ID) methodology, an applied qualitative approach designed to generate practice-relevant knowledge in health research. We purposively sampled 20 stakeholders representing diverse areas of expertise and geographic regions across World Bank country income classifications. We conducted, video-recorded, and transcribed in-depth, semi-structured interviews. Using constant comparative analysis (CCA), we identified recurring, convergent, and contradictory patterns.

**Results:** The analysis identified five overarching themes. The first two, exploratory themes, included: *Patient-Centred Understanding of Retention in HIV Care,* which captured how stakeholders conceptualized retention in their respective contexts, and *Operationalization of Retention Measures,* which explored the key components used to measure retention. The next two, explanatory themes, included *Purpose-Driven Definitions of Retention*, which described how retention measures were selected based on their intended use; and *Building Capacity through Shared Understanding and Integrated Action*, which emphasized retention as a cyclical, interconnected process dependent on collaboration between patients and health systems. The final, prescriptive theme, *Advancements Shaping Retention*, reflected stakeholders’ shared vision of improving retention through innovations in HIV treatment and technology.

**Conclusions:** The findings suggest that stakeholders operationalize retention measures in line with specific objectives and individual health goals, while remaining attentive to contextual realities. Retention measures should remain flexible and patient-centred, rather than relying on a single rigid standard.

## Background

HIV remains a global public health challenge, with an estimated 40.8 million people living with HIV (PLHIV) in 2024. Despite expanded access to antiretroviral therapy (ART), the global burden of new HIV infections and AIDS-related deaths remains high (1). To address these gaps, the World Health Organization (WHO) has prioritized retention in HIV care as a cornerstone of its global strategy to end the epidemic by 2030 (2, 3).

WHO defines retention as “the regular attendance of individuals at HIV care services based on their clinical needs.” (4). Retention in HIV care promotes adherence to ART, supports viral load suppression, and reduces HIV transmission, directly improving HIV-related health outcomes (5, 6). Beyond individual outcomes, retention strengthens health systems, guides program and policy planning, and optimizes resource allocation (7–9). Yet translating these benefits into practice is challenging, as retention in HIV care is a complex, dynamic, and multidimensional process shaped by individual, social, and contextual factors across the HIV care cascade (10–14). The influence of these factors is evident in current global estimates: the WHO’s 2025 Global and Regional HIV Statistics Report indicates that average retention in HIV care globally is 77% (range 62–90%), with 73% (65–82%) of those retained achieving virological suppression as of 2024 (1).

The WHO definition provides limited operational guidance for end-users due to its broad scope and vague terminology, thereby limiting evaluation, comparability, and evidence-informed decision-making across programmes and settings (4, 15, 16). Heterogeneity in denominators in retention measures has led experts to question the validity of retention estimates (16, 17).

Researchers have long struggled to define a universally accepted measure, testing multiple approaches in pursuit of consensus (18, 19). While observational studies comparing retention measures within the same population have demonstrated correlations among measures, no single measure has proven clearly superior (18–28). Systematic reviews have catalogued a wide range of definitions, each with specific strengths and limitations. However, the heterogeneity among these measures complicates selecting a single, definitive measure (29–31).

Building on these efforts, some researchers have applied qualitative methods and consensus-based processes to develop context-specific measures that align with local realities (32–35). National and organizational initiatives also exist. For instance, the Centers for Disease Control and Prevention (CDC) in United States, and the Ontario HIV Treatment Network (OHTN) in Ontario, Canada, track viral load as a proxy for retention, since suppressed viral load indicates adherence to ART (36–38). However, viral load testing alone is not a reliable indicator of retention due to limitations in laboratory capacity, test accuracy, patient discomfort, and data linkages across services (39–41). Consequently, consensus on a “gold standard” measure of retention remains elusive (16, 19, 29, 30).

To address the ongoing controversy over retention measurement, this study explored the experiences and perspectives of HIV stakeholders on how they define and measure retention in HIV care within their own contexts. Specifically, it investigated how HIV stakeholders interpret and operationalize retention in clinical practice and research (42). Our research question asked: What are the perspectives of HIV stakeholders on measuring and understanding retention within their specific contexts?

## Methods

### Design

We adopted an interpretivist paradigm that views retention in HIV care as co-constructed through the diverse, subjective experiences of HIV stakeholders. We used Interpretive Description (ID), a robust and flexible qualitative methodology designed for complex questions in applied health research, to guide sampling, data collection, analysis, and attention to rigour. ID responds to clinical variation and identifies patterns in real-world contexts, generating context-sensitive and clinically actionable knowledge that informs evidence-based decision-making in both clinical and policy settings. It emphasizes practical understanding over abstract theory (43).

### Participant recruitment

For this study, we recruited experienced HIV stakeholders worldwide. We categorized HIV stakeholders in three groups based on their primary roles in the HIV response: policy development (policymakers, clinical guideline developers), health research (researchers and funding bodies), and clinical practice (HIV clinicians, public health professionals, and PLHIV)(44). We applied multiple purposive sampling techniques to ensure representativeness across areas of expertise, and geographic regions based on the World Bank Country Income Classifications: low-income (LIC), lower-middle-income (LMIC), upper-middle-income (UMIC), and high-income countries (HIC) (45). We used criterion sampling to select participants who had authored at least one randomized controlled trial (RCT) on HIV retention or had at least five years of experience in HIV care. We then applied maximum variation sampling to maximize diversity across stakeholder groups and income categories. Finally, we used respondent-driven sampling to allow participants to refer additional experts, expanding the original pool (46).

We identified the corresponding authors of 60 published RCTs on HIV care retention from the <u>CASCADE database</u> (47). Of these, 15 were contacted, and six responded, and an additional eight HIV care experts were recruited through a respondent-driven sampling approach. To recruit patient representatives, we publicized our study through an advertisement shared with HIV-focused organizations (e.g., Realize: The National HIV/AIDS Resource Center [51]) and online patient communities (e.g., POZ Community Forum [52], the Global Network of People Living with HIV [GNP+] [53]), ultimately recruiting six patient representatives [51, 52]. Recruitment continued until a representative sample across stakeholder groups and geographical regions was achieved, ensuring sufficient information to address the research question (48). Four members of the study team (NR, JX, ME, GR) contacted potential participants via email and sent follow-up messages to non-respondents (see Appendix 1). Participants who agreed to join the study received detailed study information and informed consent forms through email. Our previously published protocol provides additional details of the recruitment process (44).

### Data collection

We conducted in-depth, semi-structured interviews using a pilot-tested guide informed by prior research on retention measures (29). The guide addressed four key areas related to retention measures and included open-ended questions to capture the full scope of the topic (49)(see Appendix 2). The interviews explored the following questions:

1. How do you define the term “retention”?
2. How do you operationalize retention in your work?
3. Which components of retention measurement do you apply in your work, and why?
4. How do you compare retention across different settings and programs?

A graduate student (NR), trained in qualitative methods conducted all interviews and debriefed with the research team after each session to review the process, evaluate data quality, and refine techniques, ensuring methodological consistency. The participants provided verbal consent at the start, and were informed of the study’s purpose, their rights, and the voluntary nature of participation. Interviews were conducted in English via a secure videoconferencing platform (Zoom Video Communications, 2023) at participants’ convenience.

### Data Management and Analysis Plan

#### Transcription

NR conducted qualitative interviews averaging 50 minutes (ranging from 35 to 100) between November 2023 and August 2025. We collected and analyzed data concurrently and iteratively, allowing early findings to inform both coding and the interview guide in real time. After each interview, three team members (JX, ME, GR) transcribed the data semi-verbatim, omitting fillers (e.g., “um,” “err”) and minimizing personal emotional content to maintain clarity, consistent with the study’s purpose of understanding participants’ perspectives on clinical practice (50). NR reviewed a random sample of five transcripts to verify accuracy. Transcripts were selected using simple randomization. We anonymized all transcripts and securely stored them on password-protected servers at McMaster University.

### Analysis and Interpretation

We imported transcripts and field notes into NVivo 15.2.2 (QSR International Pty Ltd, 2024) and applied an inductive, data-driven coding approach. Members of the study team (NR, JX, ME, GR) independently coded the transcripts using a modified constant comparative analysis (CCA). Using CCA, we categorized data, compared cases, and identified patterns, variations, and emerging concepts in retention in care (51). The analysis unfolded iteratively in three steps. First, we conducted line-by-line open coding to break down the data into discrete meaning units, generating concepts and uncovering novel theoretical insights. We refined codes iteratively alongside data collection until sufficient information power was achieved (48). Second, axial coding grouped related codes into categories, identified relationships, and formed overarching themes and subthemes. Finally, selective coding integrated categories into core themes, constructing a coherent interpretive framework to explain variation in retention measure (51–53).

The team members met with the lead investigator (AJ) approximately every two to three interviews to review emerging themes and reach consensus. They constructed a coding tree, examined interrelationships, and ensured diversity in expertise and geography. Findings are illustrated using participant quotes, identified by participant number in parentheses.

### Reflexivity

The lead author (NR), an Asian female doctoral student whose research focuses on HIV care cascade, is not a clinician and does not work directly with PLHIV, which positioned her as an outsider to the clinical realities of HIV care. At the same time, her disciplinary training and prior work on retention measures provided an insider perspective that informed the study design, development of the interview guide, and the interpretation of findings. She approached this study from a post-positivist paradigm, shaped in part by her prior experience with the challenges of comparing and synthesizing evidence across studies because of heterogeneity in retention measures (Rehman et al., 2024) (29). Acknowledging the influence of her positionality, NR maintained a reflexive journal documenting assumptions, methodological decisions, contextual observations, and responses to participant interactions. This process enhanced transparency and ensured that participants’ perspectives remained central to meaning-making, consistent with the principles of Interpretive Description (53).

### Trustworthiness

The study followed Lincoln and Guba’s framework to achieve rigor and trustworthiness, highlighting credibility, transferability, dependability, and confirmability (54). We enhanced transferability by recruiting a diverse sample and providing rich contextual descriptions. We strengthened credibility through prolonged engagement, iterative analysis, independent coding, peer debriefing, expert consultation, and member checking. We maintained dependability by applying systematic coding procedures, detailed memoing, and logic maps to ensure consistency and transparency. We reinforced confirmability by creating an audit trail that documented analytic decisions.

## Results

### Participant demographics

The final sample comprised 20 HIV stakeholders, 12 (60%) of whom identified as female. Eleven of the participants (11/20, 55%) were affiliated with high-income countries, and four of these reported collaborations in low-income countries (4/11, 36%). Participants often held multiple roles across categories. The largest group within these overlapping roles consisted of researchers from high-income countries (10/20, 50%). Table 1 summarizes participant characteristics.

**Table 1.** Characteristics of Participants (n = 20)

| Participant ID | World Bank Country Income* | Role (s) | Gender |
| --- | --- | --- | --- |
| P01 | HIC<sup>a</sup>/LIC<sup>b</sup> | HIV specialist; Researcher | Female |
| P02 | HIC | Patient; Activist | Male |
| P03 | HIC | HIV specialist; Researcher; Policy | Female |
| P04 | HIC | HIV specialist; Researcher | Male |
| P05 | HIC/LIC | HIV specialist; Researcher | Male |
| P06 | HIC | Patient; Activist | Male |
| P07 | HIC | Patient | Male |
| P08 | HIC | Patient | Male |
| P09 | HIC | Patient | Male |
| P10 | HIC | Patient | Female |
| P11 | HIC | HIV specialist; Researcher | Female |
| P12 | HIC | Patient | Female |
| P13 | UMIC<sup>c</sup> | HIV specialist; Researcher; Policy | Female |
| P14 | UMIC | Researcher | Female |
| P15 | UMIC<sup>d</sup> | HIV specialist; Researcher | Female |
| P16 | UMIC | HIV specialist; National program leader | Male |
| P17 | HIC/LIC | HIV specialist; Researcher | Female |
| P18 | HIC | Researcher | Female |
| P19 | HIC/UMIC | HIV specialist; Researcher | Female |

| P20 | HIC/LIC | Researcher | Female |
| --- | --- | --- | --- |

The analysis identified five overarching themes. The first two were exploratory: *Patient-Centred Understanding of Retention in HIV Care,* which captured how stakeholders conceptualized retention within their respective contexts, and *Operationalization of Retention Measures*, which explored the key components used to measure retention. The next two themes were explanatory: *Purpose Shapes the Definition of Retention*, which described how retention measures were selected based on their intended use, and *Building Capacity through Shared Understanding and Integrated Action*, which emphasized retention as a cyclical and interconnected process dependent on collaboration between patients and health systems. The final, prescriptive theme, *Advancements Shaping Retention*, reflected stakeholders’ vision of improving retention through advances in HIV treatment and technology. The most frequent coded data segments pertained to contextual (n = 144) and individual disparities (n = 88). Figure 1 presents the final themes, Appendix 3 lists supporting quotes, and Appendix 4 provides coding frequencies.

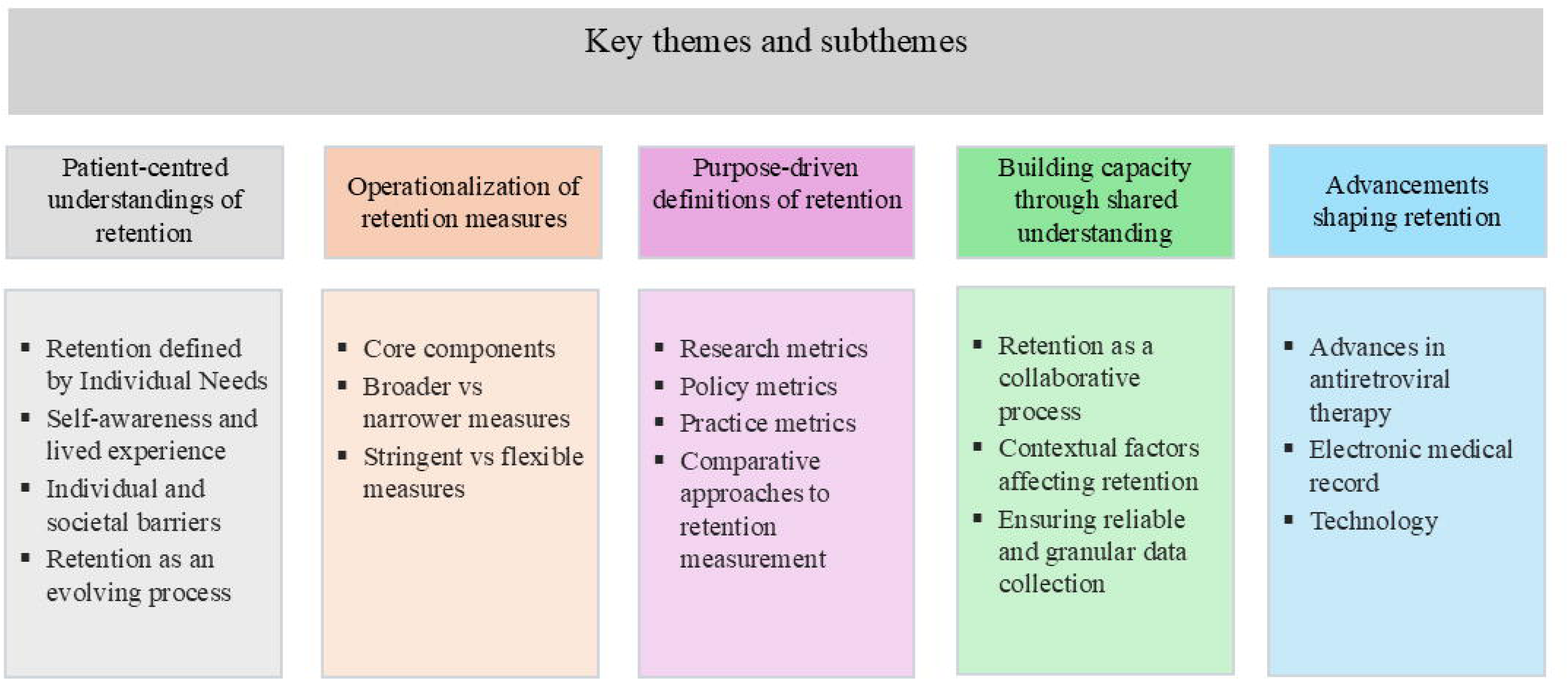

### Theme 1: Patient-Centred Understandings of Retention

Participants described retention in HIV care as a multidimensional, dynamic, and lifelong process essential for optimizing health outcomes. They emphasized that retention extends beyond adherence to HIV-specific treatments and requires a holistic understanding of well-being, including socio-economic and structural factors, highlighting the need for ongoing monitoring.

“Comprehensive medical care focuses on both HIV treatments and factors such as social determinants of health, housing, food, and stability and security. It also includes mental health conditions and substance use. To me, comprehensive is inclusive of not just HIV-specific treatment…it’s about the other factors in someone’s life that can influence how they manage their HIV infection (P04).”

#### Subtheme 1: Retention Defined by Individual Needs

Participants emphasized that retention in HIV care is inherently dependent on the individual needs of each patient. Being “retained” was understood relative to each person’s unique circumstances and adherence to their own personalized treatment plan.

#### Subtheme 2: Self-Awareness and long-Term Experience

Participants described retention in HIV care as an intentional, evolving process dependent on self-management and engagement. They noted that “retention” can be ambiguous, with some preferring “adherence to care” to emphasize patients’ active role (P06). Many noted that PLHIV do not consider visit frequency a key measure of care, leading those with limited awareness to prioritize immediate needs over long-term care. As self-awareness develops, PLHIV gradually become “experts” in managing their own health (P20). PLHIV who remain in care tend to focus on achieving meaningful health outcomes, viewing clinic appointments and laboratory tests as integral components to the process. Participants’ personal perspectives strongly shaped their experience of retention, with some describing the care process itself as therapeutic. Across narratives, lack of knowledge and self-awareness emerged as barriers to sustained retention. Participants advocated for a more comprehensive understanding of retention, one that integrates education, self-awareness, and provider support to better link lived experiences with health outcomes. One patient participant volunteered in a Ryan White HIV/AIDS Program quality improvement campaign and reported “the campaign increased understanding of retention measures and contributed to a 30–40 % improvement of retention across providers” (P02). Figure 2 presents the campaign poster.

**Figure 2.**
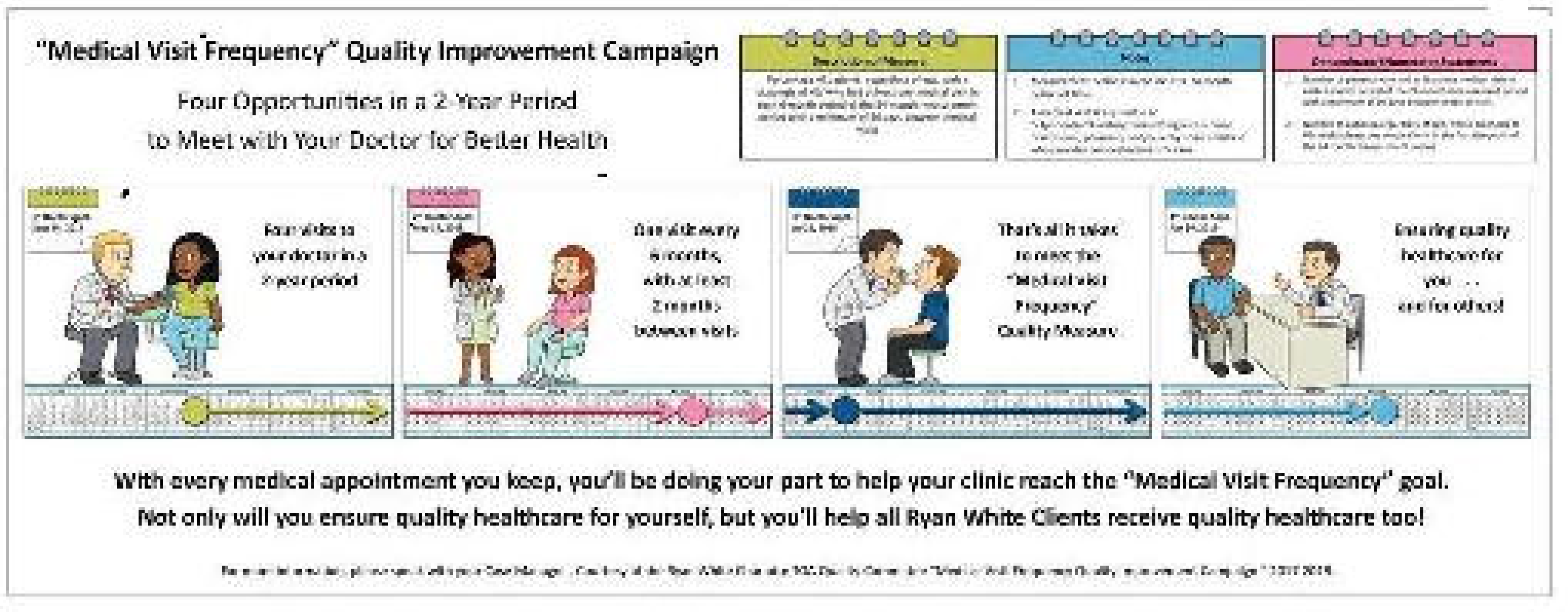
Ryan White HIV/AIDS Program quality improvement campaign poster focused on retention in HIV care.

#### Subtheme 3: Individual and Societal Barriers Determining Retention in Care

Participants described retention in HIV care as being shaped by multiple, intersecting factors at individual, clinical, and societal levels. Clinicians specifically highlighted the heterogeneity of patient needs, including challenges in distinguishing HIV-related visits from those for other health concerns.

Individual-level barriers varied between newly diagnosed patients and those who had been stabilized in care. Early challenges included adjustment to medication routines, acceptance of the diagnosis, disclosure, particularly in sero-discordant relationships, and trust-building with providers, with some participants noting that patients initially focused on symptomatic relief rather than quality of life or prevention. Other individual-level barriers leading to sup-optimal retention in HIV care included substance use, smoking, depression, comorbidities such as diabetes, treatment burdens such as blood tests and medication side effects, and age-specific needs, ranging from pediatric formulations and adolescent service transitions to older adults’ comorbidities.

Societal and structural factors that make access to care difficult include stigma, discrimination, physician burden, unstable housing, food insecurity, work schedules, caregiving responsibilities, and transportation challenges. Additional structural considerations included immigrant status, language barriers, education level, family dynamics, and disclosure of HIV status.

#### Subtheme 4: Retention as a Temporal and Evolving Process

Participants recognized retention as a lifelong process, noting that it unfolds over time rather than existing as a static state. Within the broader context of engagement, they described retention as dynamic and shaped by life circumstances. Some preferred to view retention as a continuous “flow,” which helps identify patients who move in and out of care as “cyclers” (P01, P19). Finally, some participants emphasized that retention is a behaviour that changes over time, rather than a fixed personal trait, shifting the focus away from labelling patients as “defaulters” or “good patients” (P17).

### Theme 2: Operationalization of retention measures

This theme explored how retention is operationalized in research and clinical practice. Participants viewed retention as “an objective measure, extending beyond subjective assessments of health based on how a patient feels or looks” (P13).

#### Subtheme 1: Core Components of Retention in HIV Care

Participants identified clinician visits as central to retention, supporting ART prescription, health monitoring, and individualized care planning. They emphasized that visit schedules should reflect patients’ health needs rather than predefined quotas. Pharmacy refills and medication possession ratios (MPR) are common proxies for retention, valued for their objectivity, ease of assessment, and cost-effectiveness. However, participants noted that these measures primarily reflect adherence to ART rather than the broader scope of HIV care. Retention estimates from these measures can be misleading due to factors such as drug shortages, dosing challenges, the risk of resistance, and arbitrary adherence thresholds (e.g., distinguishing between 79% and 80% adherence). Viral load was viewed as a surrogate indicator of retention, reflecting the goals of retention in care and virological suppression, but it captures only part of the picture, excluding factors such as visit frequency, encounter types, and the resources required to achieve suppression.

#### Subtheme 2: Broader vs Narrower Measures

Participants highlighted the distinction between measuring retention narrowly, using a single “touch point” (e.g., clinic visits, lab tests, medication collection), and measuring it more broadly by combining “multiple touchpoints” (P01, P19). These touchpoints are interconnected and often overlap; however, they capture different aspects of retention. The selection of the core components has important implications for measurement and interpretation. Broader, composite measures were seen as more comprehensive, reflecting the multifaceted nature of care, while aligning with patients’ lived experiences. However, participants, specifically researchers, noted that such measures pose challenges in data collection and comparability. Narrower definitions, while less comprehensive, were valued for their practicality and ease of routine monitoring across different settings.

#### Subtheme 3: Stringent vs Flexible Measures

Participants emphasized that retention should accommodate diverse patient needs rather than rely on rigid visit schedules. They favoured flexible time windows for clinical touchpoints to avoid misclassifying minor delays as non-retention. This approach aligns with the concept of “minimally acceptable care,” in which patients are considered retained if they maintain optimal medication adherence, remain virally suppressed, and attend at least one visit annually (P05). Participants noted the tension between stringency and flexibility: overly strict criteria risk misclassifying stable patients, while excessive flexibility may obscure early warning signs, such as “cyclers” who intermittently disengage (P01). They highlighted the importance of longer observation periods to capture key fluctuations in care.

### Theme Three: Purpose-Driven Definitions of Retention

Participants emphasized that definitions of retention should be purpose-driven, with measures tailored to their intended use in research, clinical care, or program evaluation. They stressed the importance of aligning measurement approaches with the underlying phenomenon being assessed, rather than merely focusing on the number or type of visits. Participants broadly accepted variation in retention definitions, reflecting their differing priorities: public health authorities track missed visits; patient advocates focus on satisfaction and well-being; administrators focus on audits; and clinicians focus on viral load, comorbidities, and preventive care. One participant summarized, “the chosen measure should answer the specific question at hand, whether it is engagement frequency or gaps in care” (P17). Participants highlighted the need for clear terminology and conceptual clarity, as well as understanding the strengths, limitations, and applicability of retention measures for specific settings and populations.

#### Sub-theme 1: Retention Measures for Research

Participants emphasized that research definitions of retention should closely align with the study’s specific purpose, ensuring that selected measures accurately capture the effects of the intervention or exposure. They noted that approaches to defining and measuring retention vary widely across studies, reflecting the importance of context-specific measures that address unique research questions and populations.

While some participants argued that research-driven definitions of retention can provide meaningful insights for policy and program development, others cautioned that such measures may have limited generalizability beyond the specific study context.

#### Subtheme 2: Retention Measures for Policy

Participants emphasized the crucial role of policy in guiding evidence-based practice, promoting cost-effectiveness, and informing public health evaluations. National policies and guidelines are often aligned with the World Health Organization’s Sustainable Development Goals (SDGs) for 2030; however, participants stressed that context is critical and that policies must be adapted to local resources and settings. Policy approaches to measuring retention were described as varying widely across low- and high-resource contexts. For example, pharmacy refill data, facilitated by integrated electronic medical records (EMRs) in public health sectors, are a feasible measure in sub-Saharan Africa, where HIV burden is high, but less practical in high-income settings, even in the United States and Canada, where EMRs are often fragmented. For comparison at national and global levels, participants noted that viral load often serves as a standard quality indicator and benchmark for service delivery and performance monitoring, rather than visit counts, largely because visit data are not consistently reportable across settings.

#### Subtheme 3: Retention Measures for Practice

Participants emphasized the need to align retention with individual health objectives in clinical practice, noting that a universal “bare minimum” does not accurately reflect patients’ real-world needs. Stable patients may require less frequent contact, whereas individuals with complex conditions need closer monitoring. Several participants advocated for personalized definitions of retention at the micro level, highlighting that retention should be guided by patient needs and clinic goals rather than rigid standards. As one participant explained, “Clinics should work on their own internal definitions and goals as providers for their patients” (P18).

#### Subtheme 4: Comparative Approaches for Retention

This subtheme underpins the rationale for initiating this work. Several participants suggested that comparisons should be based on retention measures as they are explicitly defined. For instance, when comparing clinical practices, they proposed anchoring the denominator of retention measures to the treating clinician’s prescribed follow-up schedule. Others suggested that authors of studies report multiple retention measures whenever feasible, enabling systematic reviewers to select the most appropriate measure for pooling. Participants emphasized, however, that reviewers must make careful and informed decisions before comparing or pooling studies to ensure a meaningful and accurate synthesis.

### Theme 4: Building Capacity through Shared Understanding and Integrated Action

Participants described building capacity as a cornerstone of effective HIV care. They emphasized that it relies on transparent communication across interconnected systems, including coordination within care teams and patients.

#### Subtheme 1: Retention as a Collaborative Process

Participants described retention in HIV care as a collaborative process that extends beyond clinic visits and requires active patient participation. They emphasized that collaboration among stakeholders at all levels, together with alignment around shared goals, forms the foundation for sustaining retention.

Participants highlighted practical strategies to foster collaboration. For instance, strategies implemented in Rwanda involve leveraging patient experts or champions to guide peers through care community outreach programs. In countries like Pakistan, where the epidemic disproportionately affects marginalized populations, such as people who inject drugs (PWIDs) and local migrants, who are difficult to trace, programs implement differentiated service delivery (DSD) models to improve outreach and engagement. Overall, participants framed optimal retention not merely as clinic attendance but as a structured, systematic, and participatory process that relies on collaboration among patients, providers, and the broader healthcare system.

#### Subtheme 2: Ensuring Reliable and Granular Data Collection

Participants emphasized that retention occurs at the individual patient level, making the collection of accurate, detailed individual data essential. They stressed that consistency in data collection matters more than the specific indicator used; whether recording scheduled, kept, or missed visits, systems must document them consistently for all patients.

In resource-limited or rural settings, retention data are often recorded manually in registers, which can be less reliable, subject to reporting delays, and difficult to share across facilities. Access to provincial or regional health data may be restricted across jurisdictions, limiting comprehensive tracking. EMRs offer an objective source of information; however, their functionality varies widely. In some low- and middle-income countries, EMRs are designed to flag patients at risk of loss to follow-up, whereas in the United States, missed visits may be removed from records if patients cancel or reschedule. Participants viewed gap scores, which quantify the duration of care interruptions, as a promising method for capturing retention over time, even though accurately calculating them in EMRs poses challenges.

#### Subtheme 3: Contextual Factors Affecting Retention

Participants stressed that retention cannot be understood in isolation from its context, and that no single global definition of retention is feasible given contextual factors such as resource availability, funding mechanisms, health system design, guidelines/policies, and broader social determinants such as poverty, stigma, and mobility. In high-income settings, retention is often defined by scheduled visits with HIV specialists and routine viral load testing. In contrast, these expectations may be unrealistic in low-resource contexts with high patient volumes and limited system capacity.

### Theme 5: Advancements Shaping Retention

Participants rejected a “one-size-fits-all” or “bare-minimum approach” to retention and referred to standardization as a “disservice to the notion” (P01, P04, P05, P17). Some noted that rigid definitions cannot account for the complex behavioural nature of retention and can obscure individual needs, while others emphasized that retention is a process outcome shaped by context. As one researcher from a low-and middle-income country explained, “If you want to have a standard definition, then healthcare delivery and context should follow similar standards globally” (P14).

Despite these challenges, participants expressed optimism about the future of HIV care. Advances in HIV research, such as the development of potent combination ART, preexposure prophylaxis (PrEP), and long acting injectables, may reduce the retention demands, highlighting the need for adaptable retention frameworks. Overall, participants viewed retention as evolving alongside these innovations, with their proper integration shaping both retention outcomes and broader HIV care delivery.

#### Subtheme 1: Advances in ART and Implications for Retention

Participants highlighted that significant advancements in ART have fundamentally transformed retention in HIV care. Early ART regimens required multiple daily medications, often causing significant side effects and a high risk of treatment failure, which necessitated frequent clinic visits. In contrast, newer combination ART regimens are more potent, better tolerated, and require less strict adherence. As a result, patients can maintain their health and feel retained even with fewer clinic visits and minimal side effects. Looking ahead to the era of long-acting injectable ART, participants were optimistic that these innovations may further shape HIV outcomes, as daily pill adherence becomes less central and optimal care increasingly depends on patients receiving their injections on schedule, highlighting the importance of retention in care in the evolving HIV care landscape.

#### Subtheme 2: EMR and Implications for Retention

Participants anticipated that the successful implementation and integration of unified EMR systems could play an increasingly vital role in ensuring consistent documentation and enabling seamless monitoring of patient retention in HIV care. However, the structure, integration, and coverage vary widely across settings. Factors such as patient mobility, system fragmentation, and geographic or political disparities often contribute to incomplete or inconsistent data collection and reporting, potentially limiting the ability to track retention in care accurately across facilities.

#### Subtheme 3: Technology and Implications for Retention

Participants are optimistic that virtual care, though relatively novel, can help overcome longstanding barriers in HIV care. Computerized platforms often include embedded features such as video interpreter services to address language barriers. Mobile health (mHealth) interventions, including automated reminders, have further facilitated adherence and retention. Digital tools such as electronic pillboxes and reminder apps enhance adherence and support patient self-management. However, participants emphasized the need for equitable access and attention to technology-related barriers across diverse populations.

## Discussion

We conducted an ID qualitative study and interviewed 20 experiential HIV stakeholders involved in HIV care to explore their perspectives on retention and its measurement in HIV care. Participants described retention as a dynamic, multilayered process shaped by contextual factors that requires adaptability to patients’ evolving needs. They emphasized maintaining high standards of care and called for robust, interoperable data systems and pragmatic retention measures tailored to context and the needs of people living with HIV (PLHIV), while acknowledging the challenges of supporting retention amid diverse professional roles and care settings.

### Strengths of the study

Guided by Interpretive Description methodology, this study explored the nuanced, context-specific dimensions of HIV care retention [48, 57]. We selected participants purposively and conducted in-depth interviews to capture diverse stakeholder insights. We applied inductive coding using a modified CCA to identify patterns across perspectives [55]. Through triangulation of diverse sources and co-construction of meaning with participants, we ensured comprehensive representation and strengthened the study’s rigour and relevance [48, 57].

### Limitations of the study

Our study included 20 participants; however, some geographic regions remained underrepresented. Despite this, our purposive sampling approach captured sufficient diversity to capture the influence of contextual factors on HIV care and retention (48). Although this work stemmed from a mixed-methods approach aimed at developing a standard measure of retention, the flexible and robust framework of Interpretive Description allowed for the inclusion of clinical variation. Rather than yielding a definitive, universally standardized measure, the findings highlight the value of embracing variation in retention measures and encourage viewing differences as strengths, rather than labelling participants as “good” or “bad.”

### Retention in HIV Care: Insights from Previous Evidence

Existing literature has predominantly defined retention in HIV care using quantifiable indicators, such as missed or kept visits and viral suppression, primarily reflecting retention in care (41, 55–57). In this study, participants similarly acknowledged the importance of these indicators but viewed retention as a complex, multidimensional phenomenon shaped by multiple factors. Consistent with the WHO’s broader view, they recognized that sustained retention is essential to ensure positive health outcomes for PLHIV (58). However, they emphasized that accurate assessment of patient health requires a comprehensive conceptual framework that captures these multiple dimensions, rather than relying solely on clinical outcomes or visit counts (17, 59, 60). Emerging patient-centred approaches, including the 10-item HIV Index (61) and the integration of health-related quality of life (HRQoL) measures [71], illustrate promising ways to assess the full spectrum of physical, mental, and social outcomes (58). Since factors influencing retention vary across populations and contexts, adapted measures remain essential to support evidence-based informed decision-making (60).

Our findings align with previous work that the phenomenon of retention in HIV care reflects the interplay between personal health needs and broader social determinants [55] and demands patient-centred and equity-oriented care frameworks [56]. When HIV strategies integrate equity, they reduce disparities and improve long-term outcomes, particularly for key populations including men who have sex with men, people who inject drugs, people in prisons or other closed settings, sex workers, and transgender people [57]. To achieve this, policies must adopt a patient-centred focus to prevent fragmentation and misaligned goals across health systems [58]. Approaches such as Health in All Policies (HiAP) and population health management foster cross-sector collaboration, enable data-driven planning, and support integrated care to address social determinants, thereby promoting holistic, equity-focused retention efforts that respond to the complex challenges faced by people living with HIV (PLHIV) [59].

Recent advances in HIV treatment and care delivery, such as the approval of long-acting injectables and the expansion of virtual care, have created new opportunities to overcome traditional barriers to retention (62, 63). However, socio-structural inequities in technology access and the availability and cost of injectables continue to limit these gains (64, 65). Retention in the evolving era of HIV care requires integrating new technologies and therapies into flexible, patient-responsive systems (15).

### Future implications

The participants in this study highlighted retention in care as a process outcome rather than an endpoint health outcome, which supports broader HIV care-related health goals (66). The study implications extend beyond HIV care, aligning with broader debates in health and social sciences on the standardization of outcome measures (67). While standardization supports comparability, tailored measures remain essential to reflect contextual and individual realities. This study guides researchers by highlighting that retention should be defined specifically for each study or setting. To support valid comparisons in the future, researchers should consider reporting multiple retention measures to capture different dimensions of care [19, 68], so end-users can select the most appropriate measure for their purpose.

Moving away from a single standardized definition of retention does not imply abandoning accountability in HIV care. Rather than outcome-based targets such as 95–95–95, retention reflects an ongoing care process that may be better supported by a set of complementary measures that capture continuity of engagement, clinical stability, and responsiveness to patient needs. This approach allows retention measurement to remain patient-centred while still supporting meaningful comparison across settings.

### Conclusion

This study highlights that retention in HIV care is a dynamic, multidimensional, and context-dependent process, requiring patient-centred, flexible, and purpose-driven measurement approaches beyond rigid standardization. By integrating contextual nuance, co-constructed knowledge, and adaptable measurement strategies, this work provides actionable insights to optimize retention and improve health outcomes across diverse HIV care settings.

## Supporting information

Supplementary_Appendix_1_COREQ

## Declarations

### Ethics

The Hamilton Integrated Research Ethics Board approved this study, including all communication protocols (HiREB #16500). All participants provided written informed consent. To protect confidentiality and ensure data integrity, we conducted interviews individually and removed all identifying information during transcription. The study adhered to the relevant ethical guidelines for research.

### Consent for publication

Not applicable.

### Data availability statement

The qualitative data generated during this study are not publicly available due to ethical restrictions and confidentiality agreements with participants. The interview guide is available as supplementary material.

### Authors’ contributions

N.R., A.J., and G.G. contributed to the study concept and design, with iterative discussions guiding the analytic approach and key methodological steps. A.J. provided overarching oversight throughout the research process. N.R. conducted the qualitative interviews and led primary data collection. N.R., J.X., M.G.E., and G.M.R. contributed to transcription and qualitative coding. N.R. wrote the original manuscript draft. G.G., L.S., J.E.H., M.M., T.P.G., D.M., J.X., M.G.E., G.M.R. and A.J. contributed to the interpretation of findings and critically revised the manuscript for important intellectual content. All authors approved the final manuscript and agreed to be accountable for all aspects of the work.

## Acknowledgment

We acknowledge Dr. Bushra Jamil, infectious diseases physician at Aga Khan University, Pakistan, and APPNA HIV MERIT leader; Dr. Catherine Orrell, HIV clinician and researcher at the Desmond Tutu HIV Centre, University of Cape Town, South Africa; and Dr. Malik Muhammad Umair, HIV Treatment Specialist, National AIDS Control Program, Pakistan, as content experts who supported this study.

We would like to thank a study participant for sharing a poster from a retention improvement campaign that enriched this work. We also thank all study participants for their time and contributions. We also thank all study participants for their time and contributions.

