## Supplementary_Appendix_1_COREQ for "Aligning Definitions with Realities: An Interpretive Descriptive Study on the Complexities of Measuring Retention in HIV Care in the Global Context": Supplementary_Appendix_1_COREQ.docx

**COREQ Guidelines reporting for Aligning Definitions with Realities: An ID study**

| **DOMAIN 1: RESEARCH TEAM AND REFLEXIVITY** | | | | |
| --- | --- | --- | --- | --- |
| **#** | | **ITEM** | **GUIDE QUESTIONS/DESCRIPTION** | **RESPONSE** |
|  |  | **PERSONAL CHARACTERISTICS** | |  |
| 1. | | Interviewer/facilitator | Which author/s conducted the interview or focus group? | Data were collected by trained and experienced qualitative researcher (NR) with support from (AJ) |
| 2. | | Credentials | What were the researcher’s credentials? E.g. PhD, MD | The researchers (HK and ZM) who led the study had MSc and PhD, respectively. All other authors’ credentials ranged from basic degrees to PhDs. |
| 3. | | Occupation | What was their occupation at the time of the study? | N.R. was a PhD Candidate in the Health Research Methods, Evidence, and Impact program at McMaster University. G.G. was an internal medicine specialist and health research methodologist at McMaster University. A.J. was an Assistant Professor in the Department of Health Research Methods, Evidence, and Impact at McMaster University. D.M. was a clinician-researcher affiliated with McMaster University and the MAGIC Evidence Ecosystem Foundation. L.S. was a researcher in global health at the Boston University School of Public Health. J.E.H. was a physician-researcher affiliated with Massachusetts General Hospital and Harvard Medical School. M.M. was a physician-researcher in infectious diseases at the University of Alabama at Birmingham. T.P.G. was a physician-researcher in internal medicine at Baylor College of Medicine. J.X., M.G.E., and G.M.R. were researchers affiliated with the Michael G. DeGroote School of Medicine at McMaster University. |
| 4. | | Gender | Was the researcher male or female? | Female (N.R., L.S., J.X., M.G.E., G.M.R., J.E.H.) and male (G.G., D.M., M.M., T.P.G., A.J.). |
| 5. | | Experience and training | What experience or training did the researcher have? | N.R. had formal training in qualitative research methods, qualitative analysis, and mixed-methods research through graduate-level coursework and supervised research. The remaining researchers had prior experience working in qualitative research, including involvement in study design, data analysis, and interpretation in health and health services research. |
| **RELATIONSHIP WITH PARTICIPANTS** | | | | |
| 6. | | Relationship established | Was a relationship established prior to study commencement? | Yes, a brief rapport was established with participants before the formal interview began. |
| 7. | | Participant knowledge of the  interviewer | What did the participants know about the researcher? e.g. personal goals, reasons for doing the research | No |
| 8. | | Interviewer characteristics | What characteristics were reported about the interviewer/ facilitator? e.g. Bias, assumptions, reasons and interests in the research topic | The interviews were conducted by N.R., who had subject-matter familiarity with HIV care and health services research, which supported informed probing during interviews. To address potential bias and assumptions arising from this familiarity, N.R. engaged in reflexive practice, including maintaining a reflexive journal throughout data collection and analysis. |
| **DOMAIN 2: STUDY DESIGN** | | | | |
| **#** | **ITEM** | | **GUIDE QUESTIONS/DESCRIPTION** | **RESPONSE** |
|  | **Interpretative descriptive methodology** | |  |  |
| 9. | Methodological orientation and Theory | | What methodological orientation was stated to underpin the study? e.g. grounded theory, discourse analysis, ethnography, phenomenology, content analysis | Interpretative descriptive methodology |
|  | **PARTICIPANT SELECTION** | |  |  |
| 10. | Sampling | | How were participants selected? e.g. purposive, convenience, consecutive, snowball | Purposive sampling across six wards to ensure diverse perspectives from key stakeholder groups (policy development (policymakers, clinical guideline developers), health research (researchers and funding bodies), and clinical practice (HIV clinicians, public health professionals, and PLHIV). We applied multiple purposive sampling techniques to ensure representativeness across areas of expertise, and geographic regions based on the World Bank Country Income Classifications: low-income (LIC), lower-middle-income (LMIC), upper-middle-income (UMIC), and high-income countries (HIC). We used criterion sampling to select participants who had authored at least one randomized controlled trial (RCT) on HIV retention or had at least five years of experience in HIV care. We then applied maximum variation sampling to maximize diversity across stakeholder groups and income categories. Finally, we used respondent-driven sampling to allow participants to refer additional experts, expanding the original pool |
| 11. | Method of approach | | How were participants approached? e.g. face-to-face, telephone, mail, email | Participants who agreed to join the study received detailed study information and informed consent forms through email. |
| 12. | Sample size | | How many participants were in the study? | We identified the corresponding authors of 60 published RCTs on HIV care retention from the CASCADE database. Of these, 15 were contacted, and six responded, and an additional eight HIV care experts were recruited through a respondent-driven sampling approach. To recruit patient representatives, we publicized our study through an advertisement shared with HIV-focused organizations (e.g., Realize: The National HIV/AIDS Resource Center) and online patient communities (e.g., POZ Community Forum, the Global Network of People Living with HIV [GNP+]), ultimately recruiting six patient representatives. Recruitment continued until a representative sample across stakeholder groups and geographical regions was achieved, ensuring sufficient information to address the research question |
| 13. | Non-participation | | How many people refused to participate or dropped out? Reasons? | Of these, 15 were contacted, and six participated. There were no dropouts. |
|  | **SETTING** | |  |  |
| 14. | Setting of data collection | | Where was the data collected? e.g. home, clinic, workplace | Interviews were conducted in English via a secure videoconferencing platform (Zoom Video Communications, 2023) at participants’ convenience. |
| 15. | Presence of non-participants | | Was anyone else present besides the participants and researchers? | No. All sessions were conducted privately. |
| 16. | Description of sample | | What are the important characteristics of the sample? e.g. demographic data, date | Detailed demographics including gender, HIV care expertise/engagement, years of experience, WHO geographical region were collected and presented in Table 1 of the manuscript. Stakeholders included people living with HIV, researchers, HIV specialist, policy makers, mothers, activist, national program leds. |
|  | **DATA COLLECTION** | |  |  |
| 17. | Interview guide | | Were questions, prompts, guides provided by the authors? Was it pilot tested? | We conducted in-depth, semi-structured interviews using a pilot-tested guide informed by prior research on retention measures (29). The guide addressed four key areas related to retention measures and included open-ended questions to capture the full scope of the topic. |
| 18. | Repeat interviews | | Were repeat interviews carried out? If yes, how many? | No repeat interviews were conducted. |
| 19. | Audio/visual recording | | Did the research use audio or visual recording to collect the data? | Yes, all sessions were audio-recorded with participants’ consent. |
| 20. | Field notes | | Were field notes made during and/or after the interview or focus group? | Yes, detailed field notes were taken to capture non-verbal cues and contextual observations. |
| 21. | Duration | | What was the duration of the interviews or focus group? | 50 minutes (ranging from 35 to 100) between November 2023 and August 2025. |
| 22. | Data saturation | | Was data saturation discussed? | Yes, data saturation was achieved when no new themes emerged across stakeholder groups. Recruitment continued until a representative sample across stakeholder groups and geographical regions was achieved, ensuring sufficient information to address the research question |
| 23. | Transcripts returned | | Were transcripts returned to participants for comment and/or correction? | No, transcripts were not returned due to, among other things logistical constraints. |
| **DOMAIN 3: ANALYSIS AND FINDINGS** | | | | |
|  | | **DATA ANALYSIS** |  |  |
| 24. | | Number of data coders | How many data coders coded the data? | Four coders (NR, JX, ME, GR) Other authors (G.G., L.S., J.E.H., M.M., T.P.G., D.M., J.X., M.G.E., G.M.R. and A.J) reviewed and commented on the analysis. |
| 25. | | Description of the coding tree | Did authors provide a description of the coding tree? | Yes, codes were generated inductively and grouped into categories and themes, as presented in Figure 1. Furthermore coding frequency is provided in appendix 3. |
| 26. | | Derivation of themes | Were themes identified in advance or derived from the data? | Themes were derived from the data through inductive coding. |
| 27. | | Software | What software, if applicable, was used to manage the data? | NVivo 15.2.2 (QSR International Pty Ltd, 2024) |
| 28. | | Participant checking | Did participants provide feedback on the findings? | Yes, summary findings were shared with selected participants. |
|  | | **REPORTING** |  |  |
| 29. | | Quotations presented | Were participant quotations presented to illustrate the themes/ findings? Was each quotation identified? e.g. participant number | Yes, quotes were used to illustrate each theme and sub-theme, however, to maintain the clarity of the knowledge they are provided separately in a supplementary table. |
| 30. | | Data and findings consistent | Was there consistency between the data presented and the findings? | Yes, thematic findings aligned with quotes and data sources. |
| 31. | | Clarity of major themes | Were major themes clearly presented in the findings? | **Yes, presented in a structured format in both narrative and table form.** |
| 32. | | Clarity of minor themes | Is there a description of diverse cases or discussion of minor themes? | Yes, variations in stakeholder perspectives were noted. |
